# Study Protocol: LIAM Mc Trial (Linking In with Advice and supports for Men impacted by Metastatic cancer)

**DOI:** 10.1101/2024.10.26.24316178

**Authors:** Brendan Noonan, Philip Bredin, Anita M. Cahill, Stephanie Corkery, Katie E. Johnston, Katarina Medved, Anne Marie Cusack, Josephine Hegarty, Mohamad M Saab, Samantha J. Cushen, Roisin Connolly, Brendan Palmer, Darren Dahly, Mike Murphy, Richard M. Bambury, Jack P. Gleeson

## Abstract

**Background:** The improved survival rate for many cancers in high-income countries demands a coordinated multidisciplinary approach to survivorship care and service provision to ensure optimal patient outcomes and quality of life. This study assesses the feasibility of introducing a Men’s Health Initiative supportive care intervention programme in Ireland.

**Methods:** This is a single-arm feasibility study involving a 12-week men’s cancer survivorship programme alongside routine follow-up care in patients with advanced genitourinary malignancies. Men with advanced/metastatic genitourinary cancer (including prostate, kidney, urothelial tract, testicular or penile cancer), are eligible to enrol, with a target of 72 participants over a 2-year period.

The intervention programme entails a twice-weekly physiotherapy-led exercise programme, along with dietetics, nursing, and psychosocial components, and regular signposting to additional available services. A Pilot Phase involving analysis of data from the first group of 6 participants to complete the programme is planned, before an Expansion Phase. Assessments will occur at baseline, on completion of the 12-week intervention, and 6 months post-intervention, and will include analyses of exercise/activity levels, body composition, muscle strength, psychological wellbeing, quality of life and resources utilised.

The primary endpoints are to determine the feasibility and acceptability of introducing a men’s cancer survivorship intervention programme into routine follow-up care in patients with advanced genitourinary malignancies. Secondary endpoints include impact of the intervention programme on quality of life, cancer-related fatigue, maintenance of weight, changes in body composition and changes in dietary intake and diet quality over the study period, as well as self-care agency and its relationship to quality of life and symptoms experienced. A process evaluation will capture the experiences of participation in the study, and the healthcare costs will be examined as part of the economic analysis.

Ethical approval was granted in November 2022, with recruitment commencing in May 2023.

**Discussion:** The programme described in this protocol provides a supportive and safe environment for the introduction of self-care interventions using a small group-based format supported by individualised counselling according to the participant’s identified needs. Findings will provide direction for the implementation of future supportive care programmes for men’s cancer survivorship care.

ClinicalTrials.gov Identifier: NCT05946993; Cancer Trials Ireland #: CTRIAL-IE 23-18; Irish Cancer Society (ICS) Study reference: MHI22BAM, UCC Sponsor Study Code: 22052.

## Background

With improved survival rates from recent cancer advancements, the long-term effects of cancer and its treatment on health and quality of life (QoL) necessitate a comprehensive patient-centred approach. Formalising a standardised survivorship pathway can effectively address persistent symptoms, support transitions through stages of the cancer journey, and empower patients to live well and thrive beyond diagnosis (1).

Genitourinary cancers, also referred to as urological cancers, in men involve cancers of the urinary system and the reproductive organs (i.e., kidney, ureteral, bladder, urethral, penile, prostate and testicular cancers) (2) and account for roughly 40% (3, 4) of all invasive cancers. Most supportive care intervention studies to date have focused exclusively on prostate cancer (5). Individually, cognitive-behavioural, telephone and web-based; physical activity/exercise-based; and rehabilitative interventions have shown great promise in improving various outcomes. This improvement, however, was often short-lived and a holistic, multicomponent approach is needed. A high prevalence of depressive symptoms, anxiety symptoms and suicidal ideation has been identified in men with prostate cancer (6), and there is strong evidence highlighting the needs of these men, particularly along the themes of illness, biographical and everyday life (7). Work and specialised support were raised specifically by younger and minority sexual orientation members respectively, while similar unmet needs were identified among men with testicular (8) and penile (9) cancers. This emphasises the urgency of appropriate and timely supportive care interventions. Such interventions are key to preventing and managing adverse effects of cancer and its treatment, reducing symptom and psychological burdens of the cancers and enhancing men’s QoL across the continuum of the cancer journey (10, 11).

This paper describes the research protocol of a study assessing the feasibility and acceptability of a comprehensive multidisciplinary intervention programme for men living with advanced/metastatic genitourinary cancers.

## Methods/design

### Study Design

“The LIAM Mc Trial – Linking In with Advice and supports for Men impacted by Metastatic cancer” is a single arm feasibility study of introducing a men’s cancer survivorship intervention programme alongside routine follow-up care in patients with advanced genitourinary malignancies. The programme entails physiotherapy, dietetics, nursing and psychosocial components, with signposting to already-available supportive care services at regular intervals and is aimed at better engaging with and supporting men in Ireland post-cancer treatment. Patients, in groups of 6, are provided with personalised care plans with supervised exercise and education sessions. There are two phases: a Pilot Phase involving the first group of 6 participants, followed by an Expansion Phase.

As an initial part of the study planning, a scoping review of 30 studies (5) was carried out to describe the effect of supportive care interventions for men with any genitourinary cancer and highlighted the lack of studies including men with non-prostate genitourinary cancers. This review informed the Pilot Phase involving 6 men with advanced prostate cancer. The initial number of participants per intervention group was determined by the Steering Group, based on the availability of the necessary exercise and measurement equipment, and with a view to promoting personalised care plans and to ensure both the safety and quality of the intervention.

The study incorporates an embedded mixed-method process evaluation informed by Medical Research Council guidance (12). This realist process evaluation addresses the context of the pilot study and factors that influence the outcomes of the study i.e. “what works, for whom, under what circumstances” (13, 14). The process evaluation will use the LOGIC model to systematically and visually present, map and share our understanding of the relationships among the resources used to underpin the intervention, the activities planned (intervention inputs, processes, actions) and outputs, outcomes, and impact (14). The key evaluation will occur at the end of the core trial activity which is a 12-week intervention programme (see Figure 1 SPIRIT Schedule and Figure 2 The Liam Mc Trial Flow Diagram)

**Figure 1.**
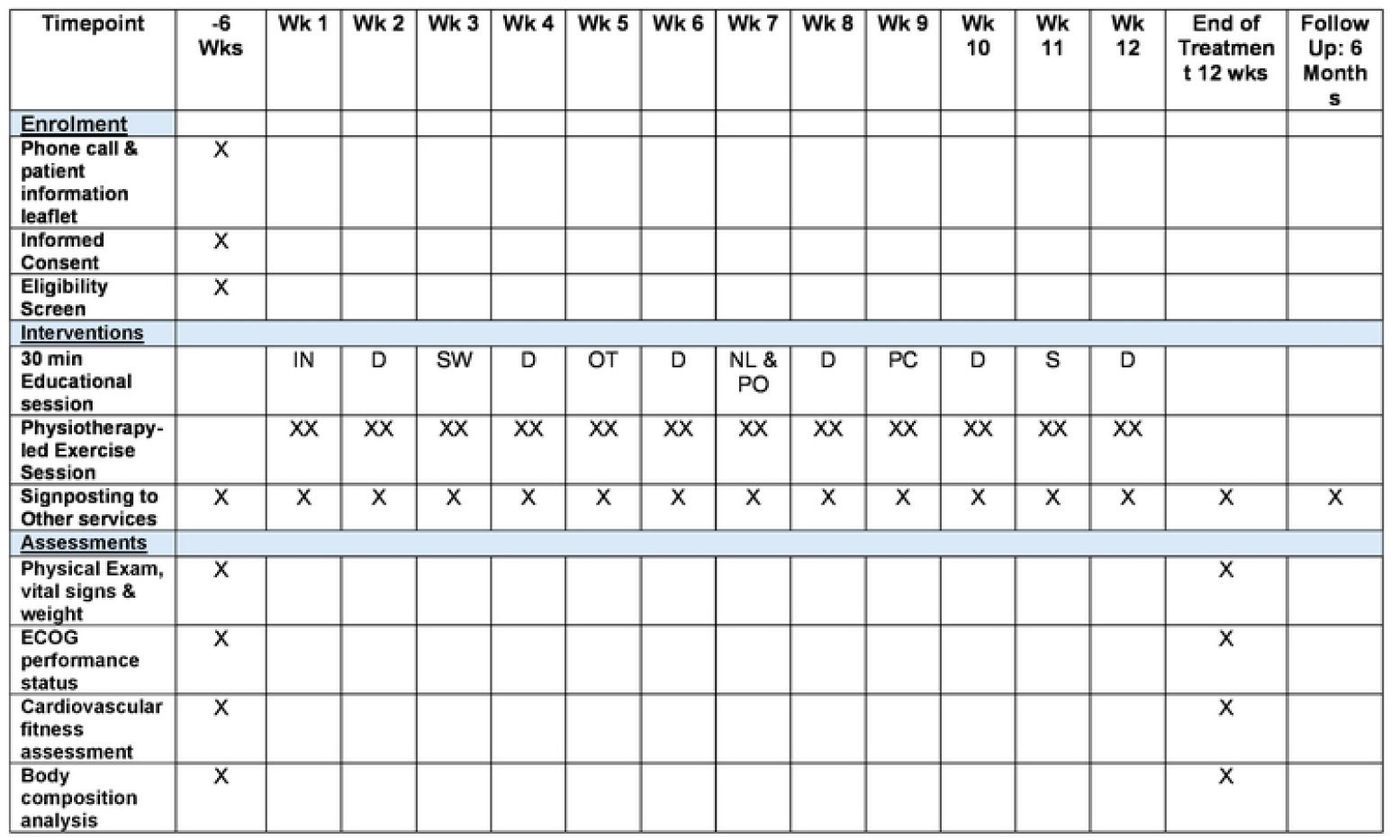
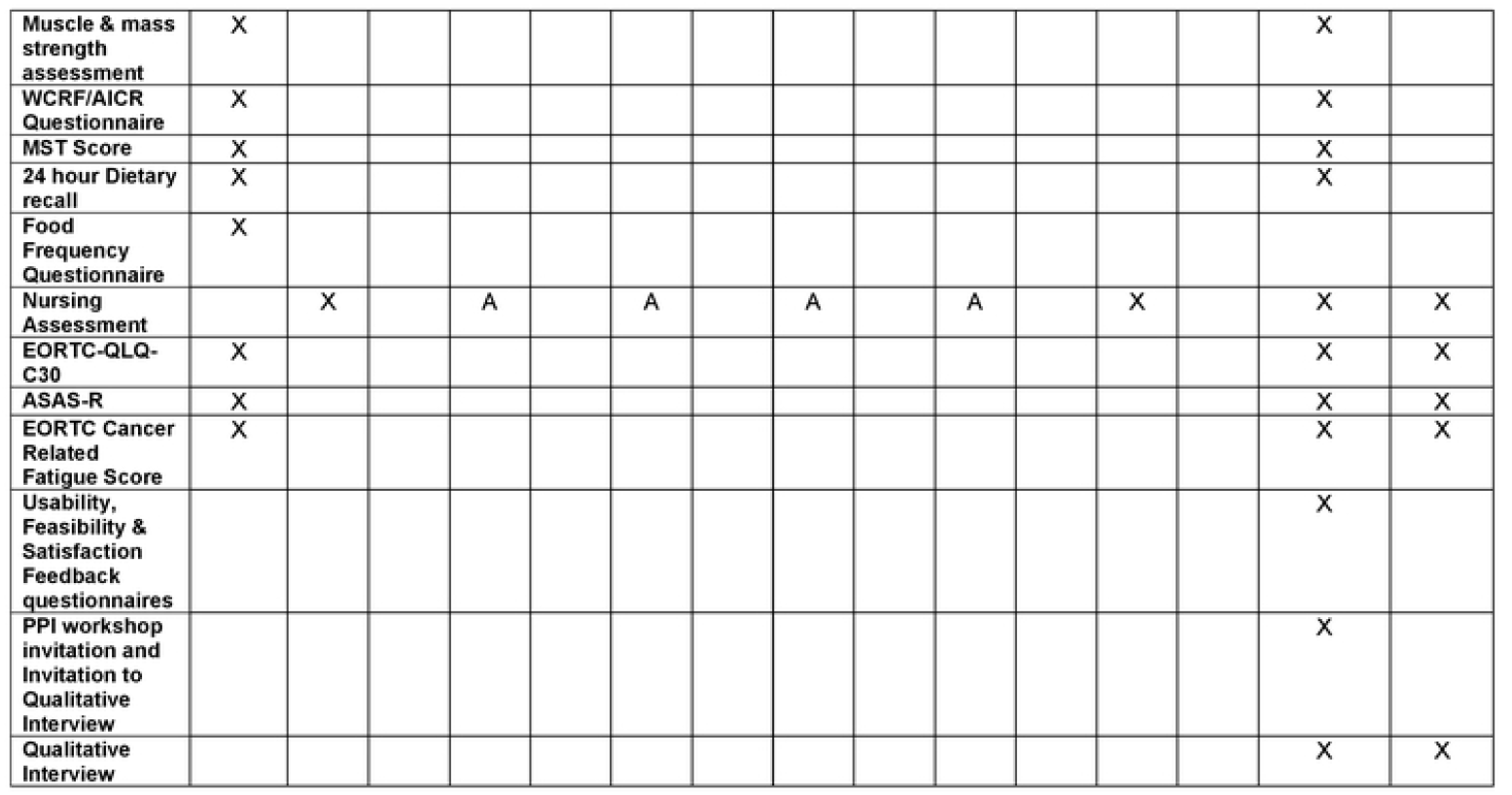
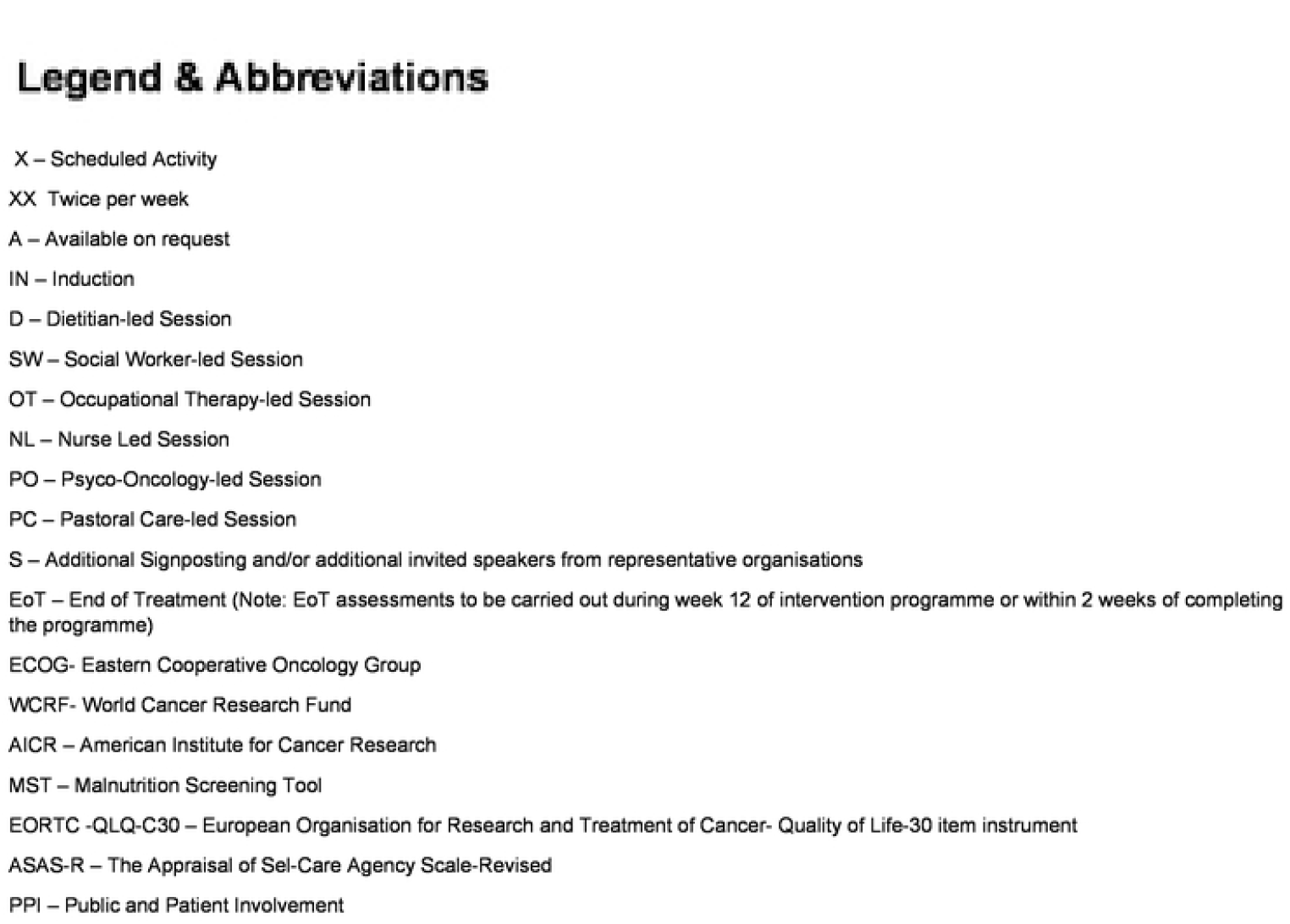
SPIRIT Schedule

**Figure 2.**
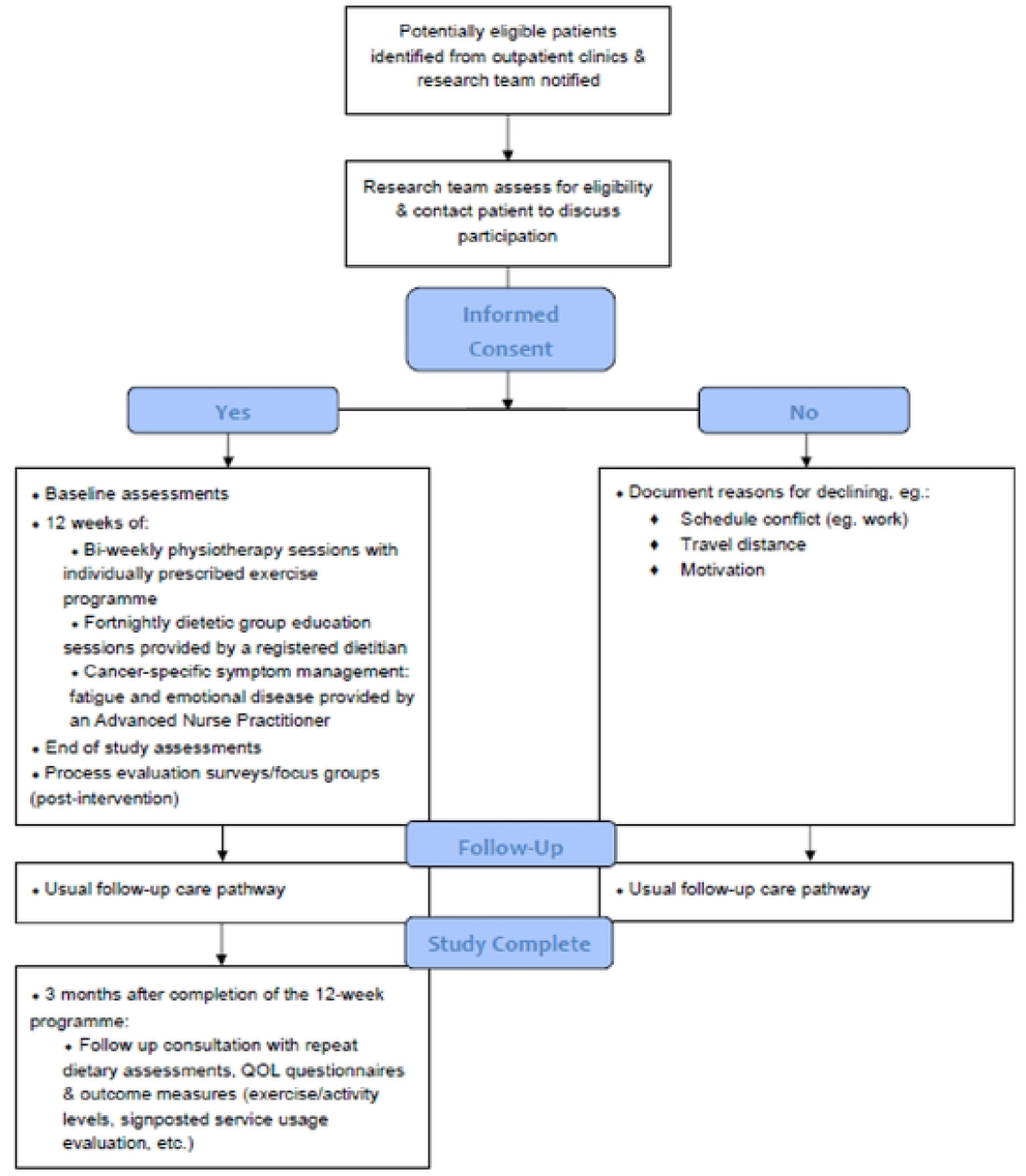
The Liam Mc Flow Diagram

### Research Ethics

This study will be conducted in accordance with the Declaration of Helsinki, the applicable sections of ICH E6 Good Clinical Practices (15) and the terms of approval of the responsible Clinical Research Ethics Committee of the University’s Teaching Hospital. Full ethical approval for the trial was granted by the Clinical Research Ethics Committee in November 2022 (ECM 4 (v) 01/11/2022). All subsequent amendments to the protocol which impacted or may impact the study’s conduct have been or will be submitted as amendments for approval to the ethics committee. The study is registered with ClinicalTrials.gov with a trial registration number of NCT05946993, Cancer Trials Ireland CTRIAL-IE 23-18 and UCC #22052. The manuscript is reported using the Standard Protocol Items: Recommendations for Interventional Trials (SPIRIT) Checklist for a clinical trial protocol (16) (Additional File 1)

### Study participants

Men aged 18 years and above, with advanced or metastatic genitourinary cancer (including prostate, kidney, urothelial tract, testicular or penile cancer), are eligible for inclusion in the study provided they are currently stable on maintenance systemic therapy, i.e. they do not have ongoing adverse events which would impact their participation in the programme at the time of commencing the 12-week intervention, or have recently completed a systemic therapy and are planned for active surveillance (i.e., no active systemic therapies at present) as part of their care for their genitourinary cancer treatment. Of note, men with resected disease (adjuvant setting) are eligible if they have commenced or completed adjuvant systemic therapy within the past 12 months and have recovered from adverse events from these treatments that would impact their participation at the time of commencing the 12-week programme.

Detailed inclusion and exclusion criteria are outlined in Table 1.

**Table 1.**
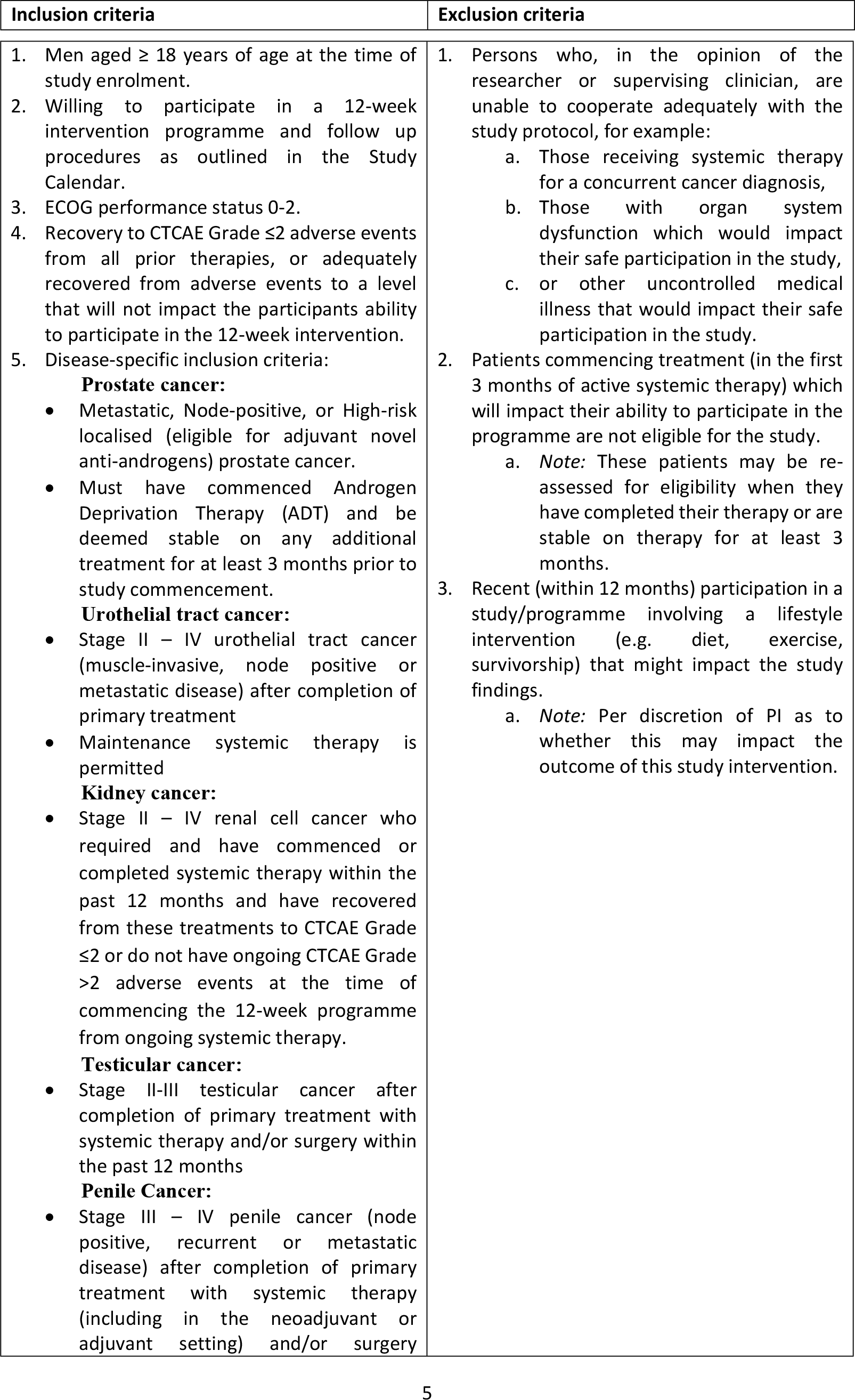
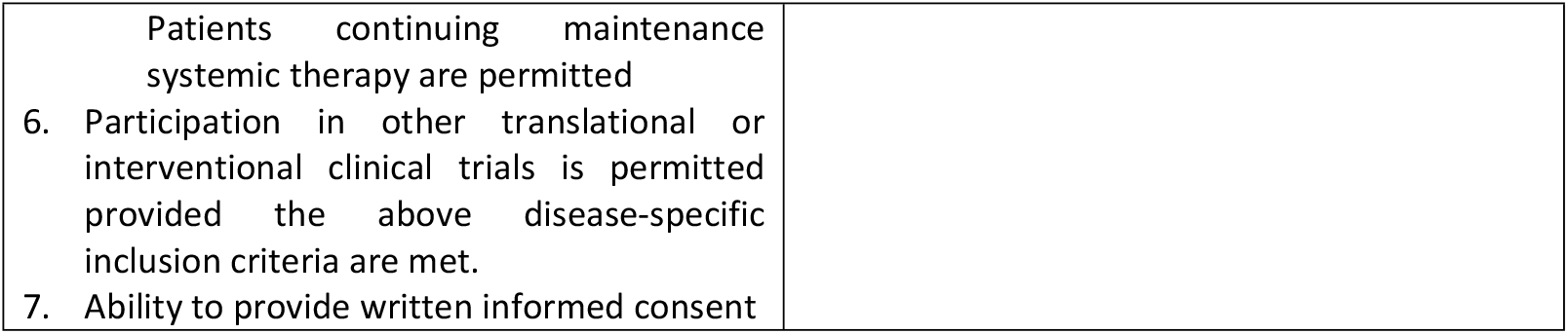
Eligibility Criteria.

| Inclusion criteria | Exclusion criteria |
| --- | --- |
| 1. Men aged $\geq 18$ years of age at the time of study enrolment.<br>2. Willing to participate in a 12-week intervention programme and follow up procedures as outlined in the Study Calendar.<br>3. ECOG performance status 0-2.<br>4. Recovery to CTCAE Grade $\leq 2$ adverse events from all prior therapies, or adequately recovered from adverse events to a level that will not impact the participants ability to participate in the 12-week intervention.<br>5. Disease-specific inclusion criteria:<br>**Prostate cancer:**<br>• Metastatic, Node-positive, or High-risk localised (eligible for adjuvant novel anti-androgens) prostate cancer.<br>• Must have commenced Androgen Deprivation Therapy (ADT) and be deemed stable on any additional treatment for at least 3 months prior to study commencement.<br>**Urothelial tract cancer:**<br>• Stage II – IV urothelial tract cancer (muscle-invasive, node positive or metastatic disease) after completion of primary treatment<br>• Maintenance systemic therapy is permitted<br>**Kidney cancer:**<br>• Stage II – IV renal cell cancer who required and have commenced or completed systemic therapy within the past 12 months and have recovered from these treatments to CTCAE Grade $\leq 2$ or do not have ongoing CTCAE Grade $>2$ adverse events at the time of commencing the 12-week programme from ongoing systemic therapy.<br>**Testicular cancer:**<br>• Stage II-III testicular cancer after completion of primary treatment with systemic therapy and/or surgery within the past 12 months<br>**Penile Cancer:**<br>• Stage III – IV penile cancer (node positive, recurrent or metastatic disease) after completion of primary treatment with systemic therapy (including in the neoadjuvant or adjuvant setting) and/or surgery | 1. Persons who, in the opinion of the researcher or supervising clinician, are unable to cooperate adequately with the study protocol, for example:<br>a. Those receiving systemic therapy for a concurrent cancer diagnosis,<br>b. Those with organ system dysfunction which would impact their safe participation in the study,<br>c. or other uncontrolled medical illness that would impact their safe participation in the study.<br>2. Patients commencing treatment (in the first 3 months of active systemic therapy) which will impact their ability to participate in the programme are not eligible for the study.<br>a. *Note:* These patients may be re-assessed for eligibility when they have completed their therapy or are stable on therapy for at least 3 months.<br>3. Recent (within 12 months) participation in a study/programme involving a lifestyle intervention (e.g. diet, exercise, survivorship) that might impact the study findings.<br>a. *Note:* Per discretion of PI as to whether this may impact the outcome of this study intervention. |
| Patients continuing maintenance systemic therapy are permitted |  |
| 6. Participation in other translational or interventional clinical trials is permitted provided the above disease-specific inclusion criteria are met. |  |
| 7. Ability to provide written informed consent |  |

### Recruitment and screening

Participants will be recruited across a university hospital group comprising three participating hospitals -Cork University Hospital, the Mercy University Hospital and the Bon Secours Hospital - within a health service region in the Republic of Ireland. A target sample of 72 participants meeting the predefined inclusion criteria will be recruited. Potential participants will be screened by the oncology clinical care team or research team to determine eligibility and all those who meet the inclusion criteria will be invited to participate. Verbal and written information regarding the study will be provided, with ample time for review before written informed consent is completed at the initial assessment. All study participants who provide written informed consent will be enrolled in the study. The Castor Electronic Data Capture (EDC) platform (https://www.castoredc.com/) will be used to collect and store the study data. Study participants will be facilitated with access to the electronic patient-reported outcome (ePRO) measurement system via Castor EDC.

Recruitment take place over a 24-month period. The study duration for each participant will involve a 12-week intervention and a follow up survey at 6 months. Strategies being taken to reach the target sample size of 72 include readily available study brochures and contact details, the use of social media and print media, and education of oncology staff in the participating hospitals about the study.

Following consent and enrolment, participants will be assigned a unique study identifier and access to the ePRO measurement system. A 6-week screening period, to assess eligibility and capture baseline assessments prior to commencing the 12-week intervention programme, will involve an initial assessment and time for consent. At the initial assessment, baseline assessments will include the following:

1. Baseline questionnaire/needs, burden of disease and quality of life assessment
2. Dietetic Evaluation
3. Physiotherapy Evaluation

Information on the questionnaires, evaluations and consent processes are contained in the trial protocol (Additional File 2).

### Intervention

The 12-week intervention programme will involve twice-weekly visits from week 1–12 (1 × 1.5-hour session, 1 × 1-hour session per week). The intervention programme will be conducted in a purposefully designed rehabilitation gymnasium. This allied health professional-led programme includes physiotherapist input twice per week, dietitian input every 2 weeks, specialist nursing input every 2 weeks, medical social worker input and psycho-oncology input, with programme oversight by medical oncologists. An outline of the programme is provided in Figure 3.

**Figure 3.**
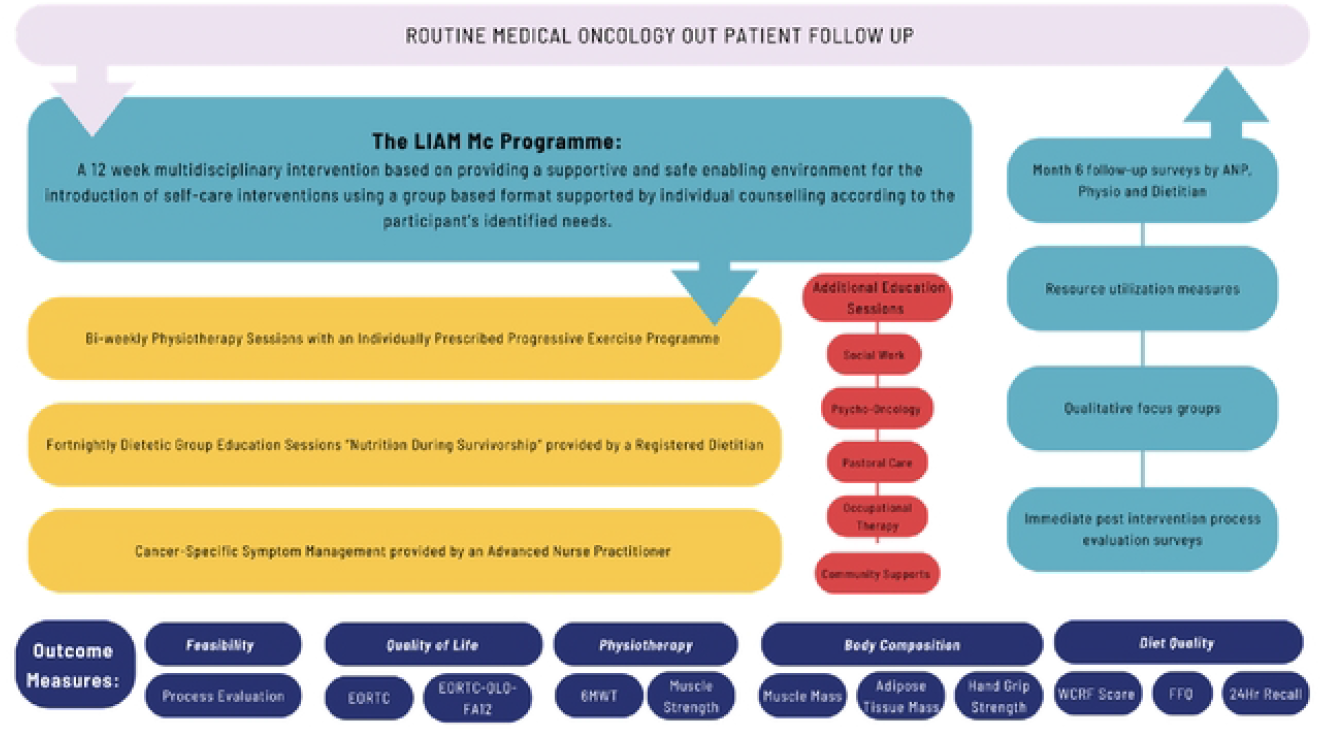
Outline of the Programme

The initial assessment visit involves assessing and managing current symptoms and needs per management pathways, referring patients to appropriate specialists, and ensuring future access to the clinic where needed during the study period. The baseline physical and nutritional assessments will be undertaken at this visit. Patients will receive individualised education on exercise, diet and symptom management plans, depending on the outcome of the baseline assessments, or according to clinical judgement. Diet education and personalised nutrition counselling will be performed throughout the study.

A study evaluation will take place, at the evaluation stage of the Pilot Phase, of potential barriers that participants perceive would prelude them in attending face to face. Anticipated barriers include access to travel, a translator, expenses incurred or family/work commitments. It is the intention of the research team to explore all feasible means to address identified barriers for later phase participants. A patient access fund is also available to help reduce barriers to enrolment. If participants cannot attend the scheduled sessions on a given day, then virtual meeting or recording options will be explored so that they can still receive the educational components of the intervention.

#### Physiotherapy

Each participant will have an individualised exercise plan developed by an oncology physiotherapist based on their medical history and scan reviews and will complete this during 2 × 1h supervised exercise sessions per week. Participants will be progressively guided through the programme, incrementally increasing in intensity or with modifications based on symptomatic presentation. This will be based on their baseline strength and cardiovascular fitness testing and grounded on evidence-based protocols previously demonstrating an effect in this patient population. A sample exercise programme is included in Additional File 3.

#### Dietetics

The program will provide both general and individualized dietary guidance to improve overall dietary quality and promote optimal body composition in participants. This includes standardized assessments, group education on diet and cancer survivorship, and personalized counselling adjusted as needed throughout the program based on individual needs and preferences identified through regular symptom surveys. Symptom surveys at baseline and 12 weeks using the malnutrition screening tool (MST) will identify nutritional risk. Patient-specific nutrition counselling will be offered to patients identified at risk. The nutrition education programme is centred around the World Cancer Research Fund’s (WCRF) recommendations for cancer survivorship (17) and prevention of non-communicable diseases.

#### Nursing & Psychosocial

This will consist of generalised discussions on areas such as erectile dysfunction, changes in masculinity, effects of hormone treatments on mood changes and body image. There will be an inclusion of practical information sessions including health systems information and managing side effects such as urinary symptoms, hot flushes, penile shortening, and loss of body hair. There will be opportunity for each man to discuss intimate concerns in a quiet and safe space.

Nursing, psycho-oncology, pastoral care and social work education sessions will inform participants on items including diagnosis shock, acceptance, coping with relationship changes with their partner and their roles, fear of uncertainty and the future, medication management, masculine identity. Psychological coaching will be offered to all participants in this study individually, to be conducted virtually for the men at a time that is convenient.

Throughout the programme, signposting of additional relevant services will also be conducted, with particular emphasis placed on these at assigned times in the schedule. At the initial assessment visit and as detailed in the study calendar, patients will be signposted to publicly available resources such as Psychosocial Coaching, Cancer Support House Cork (ARC House), Pharmacist Medication Review, Nursing Review, Medical Review, Social Work Supports, Psycho-Oncology Supports.

### Participant flow through the study

Figure 2 depicts an overview of study participants using the Consolidated Standards of Reporting Trials (CONSORT) flow diagram (18). The Pilot Phase 12-week programme of the first 6 men will be completed and analysed prior to commencement of the Expansion Phase from Q3 of year 1 onwards. Patients will participate in the programme for a total of up to 12 months. They will complete surveys during the first 12 weeks of the study (Screening/Baseline and End of Treatment) and then at the 6-month timepoint. A focus group with study participants will take place at the end of the 12-week programme. In addition, participants will be invited to Public and Patient Involvement (PPI) engagement meetings quarterly after completion of the 12-week intervention programme and for the duration of the study period (2 years).

Follow up for routine medical oncology care and surveillance will take place as normal, through the medical oncology clinics, see Figure 3. Study-specific follow up will take place 6 months after completion of the 12-week intervention programme.

If the participant decides to cease involvement, the research nurse will liaise with the participant to discuss their reasons for not continuing in the study. Reasons for declining participation in the study or discontinuation during the study period will be collected as part of the feasibility outcomes. Specific reasons for withdrawal of interest will be recorded in the relevant case report form.

### Study objectives

The primary objective of the study is to demonstrate the feasibility of introducing a men’s cancer survivorship intervention programme alongside routine follow up care in patients with advanced genitourinary malignancies. Secondary objectives include evaluation of the impact of the intervention programme on quality of life, cancer-related fatigue, body weight, lean tissue mass, fat mass, muscle strength and nutritional risk, dietary intake and quality, self-care agency, resource utilisation of signposted services and economic impact of a men’s cancer survivorship intervention programme.

### Study Outcomes/Measures

Data collection will occur at three designated timepoints; Initial assessment, after completing the 12-week intervention and at 6 months post intervention. Completion of a process evaluation as part of the feasibility testing and secondary outcome measurement will also occur after all participants have completed the 12-week intervention.

#### Primary Study Endpoint

The primary endpoints of this study are the feasibility and acceptability of introducing a men’s cancer survivorship intervention programme into routine follow up care in patients with advanced genitourinary malignancies.

The programme acceptability will be measured with each participant at programme completion and at the 6-month timepoint. PPI feedback in the focus group sessions, at quarterly engagement meetings will also contribute to understanding the level of acceptability of and support for the intervention. Determination of feasibility is multifactorial, and this will be reviewed by the Steering Committee throughout the project and during the scheduled PPI engagement meetings to inform subsequent phases. Qualitative focus groups where participant feedback is provided, and the final analysis by the steering and research groups in conjunction with key stakeholders, will all contribute to the final determination of the feasibility of the programme. This will help explore what elements of the intervention worked, what elements did not work, and what elements require improvement. A purposefully-designed brief feasibility questionnaire will be used in line with previously developed questionnaires by our team (19).

#### Secondary Study Endpoints

Impact of the intervention programme on quality of life will be measured with the European Organization for Research and Treatment of Cancer Quality of Life Questionnaire (EORTC QLQ) C30 (20), quality-adjusted life-year (QALY) (EQ-5D-5L) (21) and Appraisal of Self-Care Agency Scale-Revised (ASAS-R) assessment (22). The impact of the intervention programme on cancer-related fatigue will be measured using a cancer-related fatigue score (EORTC QLQ-FA12). The impact of the physiotherapy and dietetic interventions will be objectively tested through maintenance of weight, changes in body composition (including lean tissue mass and fat mass), muscle strength, physical function, and cardiovascular fitness; and changes in dietary intake and diet quality over the study period resulting from the dietetic intervention. Body composition will be assessed using bioelectrical impedance analysis and ultrasound. Dietary assessments will include two 24-Hour Dietary Recalls and the WCRF dietary quality score at initial assessment and completion of the programme (12 weeks) and a Food Frequency Questionnaire (FFQ) at initial assessment and at 6-month follow up. Self-care agency and its relationship to quality of life and symptoms experienced will be measured using the ASAS-R.

The number of signposted resources used will be recorded. Satisfaction of participants and Health Care Professionals with the programme and their perceptions of the system’s usability will be recorded via a Usability, Satisfaction and Feasibility Questionnaire. Participants, health care professionals and the broader team involved in the development and implementation of the programme will be invited to provide feedback after the completion of their involvement in the study through qualitative interviews

### Adverse events

Adverse events (AEs) of interest (those deemed potentially related to the intervention) will be recorded and assessed for relationship to the intervention programme throughout the study period. Adverse Events will be documented and graded in accordance with Common Terminology Criteria for Adverse Events (CTCAE) v5.0. Patient reporting of an AE to a health care professional will trigger recording of the AE.

Serious Adverse Events (SAEs), defined for the purposes of this study as CTCAE v5.0 grade ≥3 adverse events of any kind, or those grade <3 but deemed by an investigator to be serious, will be reported to the LIAM Mc Steering Group for review at the following meeting. SAEs of grade ≥4 will trigger a pause in the study for safety reasons until the SAE has been formally assessed and a decision made to either proceed with or terminate the study.

The research team do not anticipate any adverse events associated with completion of the patient-reported outcomes. Patients will have the option of skipping questions they do not wish to answer. In the event that study participation does result in any event that has significant negative consequences for the subject, this will be recorded by an investigator in the EDC (Electronic Data Capture) and will be reported to the Sponsor and Ethics committee.

### Data management and analysis

The study’s protocol is accompanied by a Data Management Plan. For confidentiality purposes, a study-specific ID code will serve as a unique identifier on the case-report forms for all study participants. The codes will be stored separately from the main study database and other study documentation. This unique ID subject code will be linked to the participant’s identifying information by a participant identity list maintained by the study’s Principal Investigator in a secure location at the clinical site of the study. The list of ID codes will not be forwarded to any third party. The purpose of the ID code is to facilitate subsequent follow-up of participants in the event of requiring symptom management intervention.

An on-site Trial Master Folder (TMF) containing confidential data in the form of hard copies of regulatory documents and participant-related information will be kept in a locked cabinet in a locked room, accessible only by the study’s research team members. An electronic TMF containing confidential regulatory documents, participant-related information, and patient public interactions will be maintained in a dedicated database in a secure location. This confidential electronic data will be stored and backed-up monthly in encrypted folders on a health service laptop, and in the validated web-based nutritional analysis software tool, Nutritics (https://www.nutrics.com/app/). Access to the database will be controlled at the level of the individual with specific roles assigned, with concomitant access rights (e.g. data manager, auditor). At the end of the study, all anonymised data will be available to the study’s principal statistician, while researchers nominated by the Principal Investigator will also be able to access the final trial dataset.

The study’s primary data catalogue, encompassing primary participant data (case-report forms) and participant responses to electronic surveys, will be collected and managed using the Castor EDC platform. This platform meets national and international guidelines with respect to data privacy and data protection standards. Once data entry is finalised, study data will be assessed for incompatible, discrepant, or clinically implausible values. Outlying values for all distributions, in isolation and over time, will be identified. Any concerning data will be reconciled against the original source data. Following the completion of cleaning, the database will be locked.

### Sample size

There is no gold standard for sample size calculation in feasibility studies (23). Given that the goal of a feasibility study is to identify problems that would impede the conduct of a larger, efficacy study, we have set the sample size at 59 based on advice from Viechtbauer et al. (24), which is aimed at being able to detect failures in study processes that would occur just 5% of the time (with 95% confidence). A target sample size of 72 allows for sample attrition (around 20% attrition rate) from men withdrawing due to illness, or waning commitment/interest. Therefore, a target sample size of 72, based on enrolling 12 groups of 6 men in Sequential Cohorts/Parallel Sampling Groups over the 2-year period, will be used and will inform feasibility of the programme.

### Statistical Considerations / Analyses

Quantitative data will be analysed using R Project for Statistical Computing and the RStudio IDE and presented using percentages, means (SD), modes, and medians with an interquartile range (IQR) as appropriate.

Qualitative data will be digitally recorded, verbatim transcripts will be prepared from the sound files, and transcripts will be checked for accuracy against the sound files and anonymised. Thematic content analysis will be used to code data in transcripts that is relevant to the process evaluation (25). NVivo software will be used to assist with the analysis. One researcher trained in qualitative research methods will analyse the qualitative data allowing for fuller immersion and to obtain an overall sense of the data. To enhance trustworthiness, this process will be checked for accuracy by a minimum of two researchers. Initial open coding will be organised into higher level coding, and thematic interpretations. Data analysis will be iterative such that early interviews can inform questions in later interviews. Quantitative and qualitative data will be integrated to provide an overall perspective on the process of implementing the intervention/study. We will do that by describing what was delivered in the intervention, what process effects were observed, then identify explanatory ‘Context + Mechanism → Process effect’ alignments that explain how the intervention, and the study more broadly, was perceived by participants, if/why this varied, and how these perceptions affected receptivity to the intervention.

Once data entry is finalised, study data will be assessed for incompatible, discrepant or clinically implausible values. Outlying values for all distributions, in isolation and over time, will be identified. Any concerning data will be reconciled against original source data. Following completion of cleaning the database will be locked.

The study sample will be described in detail. Continuous variables will be described by their means and SDs, medians and IQRs, and their range; while categorical variables will be described by their counts and percentages in each category.

Feasibility outcomes will be similarly described. These include the number of enrolled patients who complete the initial assessment and follow-up assessments, and the number of patients who partake in all planned activities versus less than all; the number of patients that require medical review and the timeframe to medical review; changes in muscle strength and mass from initial assessment to end of programme assessments; changes in dietetic assessments from initial assessment to end of programme assessments; changes in QOL outcome measures from initial assessment to end of programme assessments; the number of patients enrolled in the programme; extra HCP time required and resources required for the intervention; and reasons for not completing the programme will be collected through Drop Out Forms and with qualitative discussions with HCPs involved in the programme.

Missing data will be evaluated, and based on what we observe, dealt with in whatever manner we find appropriate based on current best practices. All analyses will be conducted using the R Project for Statistical Computing and the RStudio IDE. All trial reporting will be done following CONSORT and the CONSORT addendum for pilot/feasibility trials (26).

### Data monitoring

The university as the study sponsor is the Data Controller responsible for the study. Castor EDC provides for a full audit trail of user actions to be recorded, and data check procedures for Castor EDC will be conducted every 3 months from the start of the recruitment process by the study team. Biannual monitoring reports will be provided to the study’s Sponsor Office, and a Clinical Trial Audit will be provided by the study’s Clinical Research Facility as required from the Sponsor Office. The study team will provide annual reports to the study’s funders as expressed in the funding agreements. Based on the nonmedical intervention and low-risk nature of participation in the study, a Data Monitoring Committee was deemed unnecessary.

### Dissemination

In consultation with the study funders, it is planned to disseminate the key findings of the feasibility trial after the end of the study whereby anonymised individual participant data sets will be shared in the form of results within peer-reviewed publications, academic conferences, workshops, and seminars. Planned public and patient dissemination outputs include public blogs, media, and outreach engagement events, including press articles and seminars. The protocol is published on the ClinicalTrials. gov website, with trial registration number NCT05946993. Post-study shared outputs including the study protocol, data dictionary, and analysis scripts will be available via the Open Science Framework.

### Public and Patient involvement (PPI)

The study team members are collaborating with PPI representatives in the design, development, implementation, analysis, and dissemination of the study. The PPI representatives include patient advocates who have had genitourinary or other cancers and represent the patient and public voice at our research meetings. PPI representatives are included in the trial steering group and will be involved in the design, conduct, analysis and dissemination of the study.

## Discussion

The aim of the LIAM Mc Trial is to assess the feasibility of a comprehensive multidisciplinary intervention programme for men living with advanced/metastatic genitourinary cancers. As part of this initiative, we seek to address the key gaps and unmet survivorship needs of men affected by cancer. The focus of this novel programme will be centred on provision of evidence to drive improvement in the survivorship supports and services for those cancers that yield a significant burden on quality of life due to morbidity related to tumour burden, local treatment effects, and/or systemic treatment effects such as androgen deprivation, for which there are still considerable challenges and resources issues (5).

The information gleaned will explain how the intervention worked and how these effects might be replicated in a Quality Improvement (QI) initiative for our patients in the future, by offering this intervention to all our patients as a standard component of clinical care. The proposed intervention strategically aligns with patient priorities (27). Its goal is to identify and manage important symptoms experienced by men impacted by effects of cancer treatment. These are outlined in Ireland’s National Cancer Strategy 2017-2026.

An important aspect of this novel programme is to demonstrate how to improve the survivorship supports and services for underserved communities of men who have not traditionally been the focus of such a formalised survivorship pathway and are recognised as experiencing disparities in terms of cancer incidence, prognosis, outcome and/or quality of life. These might include, for example, members of the Travelling community, the LGBTQ+ community, ethnic minority and migrant communities, communities with social disadvantage and/or socio-economic challenges, or specific mental health issues likely to impact their ability to have a positive outcome from a cancer diagnosis.

The survivorship programme is a 12-week group-based intervention programme for men with advanced cancer, encompassing intensive multidisciplinary input to provide men with personalised tools and coping mechanisms for life with cancer. The programme is based upon providing a supportive and safe enabling environment for the introduction of self-care interventions using a group-based format supported by individualised counselling according to the participant’s identified needs.

While efforts have been made to reduce the risk of confounding factors impacting the study results, a number of limitations will need to be considered in terms of the study findings. Firstly, since the study is a single arm trial, it is by design not possible to determine differences in symptoms and quality of life outcomes between the study group and the general population of men who are survivors of metastatic cancer undergoing standard of care treatment. Secondly, inclusion criteria (for logistical reasons) means that participants with a cancer diagnosis from a site other than the genitourinary system are excluded from the study. Thirdly, outcome measurement in a complex environment such as this has a risk of confounding factors, for example, participant experience will likely relate to their interactions with the healthcare practitioners providing the intervention. Fourth, the burden introduced by the FFQ dietary assessment and other questionnaires may reduce the acceptability of the intervention to participants. A weakness of the FFQ is that it captures the previous 12-month dietary intake which can be significantly altered due to treatment and side effects and the accuracy of reporting food frequencies and portions requires good participant memory, literacy, and numerical skills.

Additional file 1. The Standard Protocol Items: Recommendations for Interventional Trials (SPIRIT) 2013 Checklist for a clinical trial protocol.

Additional file 2. Clinical trial protocol

Additional file 3. Sample exercise programme

## Data Availability

No datasets were generated or analysed during the current study. All relevant data from this study will be made available upon study completion.

## Acknowledgements

The development of a men’s cancer survivorship intervention programme within the HSE South/Southwest hospital group is a collaboration between the Irish Cancer Society, Cork University Hospital, UCC Cancer Trials Group (CTG), the Enhancing Cancer Awareness and Survivorship Programmes (ECASP) at the School of Nursing and Midwifery UCC; and regional Cancer Support services including ARC House, Daffodil Centres and Recovery Haven amongst others. The Authors would like to thank the Mardyke Arena UCC staff for the provision of excellent facilities, and the staff of the Cardio Rehab gym in CUH. Special thanks also to our PPI steering group representatives Mr Martin O’Sullivan, Mr Gerard Ingoldsby and Mr Alan Gaine and to Dr Fahmi Ishmail, Ms Aisling Deasy, Dr Daniel Nuzum, Mr Livingstone Kiwanuka, Ms Sinead Power, and Ms Jane Prendergast for their fantastic contributions to the study for the benefit of the participants.

## References

1. Hollen PJ, Hobbie WL. Establishing comprehensive specialty follow-up clinics for long-term survivors of cancer. Providing systematic physiological and psychosocial support. Support Care Cancer. 1995;3(1):40–4.

2. Dy GW, Gore JL, Forouzanfar MH, Naghavi M, Fitzmaurice C. Global Burden of Urologic Cancers, 1990-2013. Eur Urol. 2017;71(3):437–46.

3. Ireland NCRi. Annual Statistical Report 2023. https://www.ncri.ie/sites/ncri/files/pubs/NCRI_AnnualStatisticalReport_2023.pdf; 2023.

4. Siegel RL, Giaquinto AN, Jemal A. Cancer statistics, 2024. CA Cancer J Clin. 2024;74(1):12–49.

5. Saab MM, McCarthy M, Murphy M, Medved K, O’Malley M, Bambury RM, et al. Supportive care interventions for men with urological cancers: a scoping review. Support Care Cancer. 2023;31(9):530.

6. Brunckhorst O, Hashemi S, Martin A, George G, Van Hemelrijck M, Dasgupta P, et al. Depression, anxiety, and suicidality in patients with prostate cancer: a systematic review and meta-analysis of observational studies. Prostate Cancer Prostatic Dis. 2021;24(2):281–9.

7. Prashar J, Schartau P, Murray E. Supportive care needs of men with prostate cancer: A systematic review update. Eur J Cancer Care (Engl). 2022;31(2):e13541.

8. Doyle R, Craft P, Turner M, Paterson C. Identifying the unmet supportive care needs of individuals affected by testicular cancer: a systematic review. J Cancer Surviv. 2024;18(2):263–87.

9. Paterson C, Primeau C, Bowker M, Jensen B, MacLennan S, Yuan Y, et al. What are the unmet supportive care needs of men affected by penile cancer? A systematic review of the empirical evidence. Eur J Oncol Nurs. 2020;48:101805.

10. Cancer MAoSCi. What is Supportive Care? https://mascc.org/what-is-supportive-care/2024 [

11. Bernat JK, Wittman DA, Hawley ST, Hamstra DA, Helfand AM, Haggstrom DA, et al. Symptom burden and information needs in prostate cancer survivors: a case for tailored long-term survivorship care. BJU Int. 2016;118(3):372–8.

12. Skivington K, Matthews L, Simpson SA, Craig P, Baird J, Blazeby JM, et al. A new framework for developing and evaluating complex interventions: update of Medical Research Council guidance. BMJ. 2021;374:2061.

13. Moore GF, Audrey S, Barker M, Bond L, Bonell C, Hardeman W, et al. Process evaluation of complex interventions: Medical Research Council guidance. BMJ. 2015;350:h1258.

14. Greenhalgh T, Wong G, Jagosh J, Greenhalgh J, Manzano A, Westhorp G, et al. Protocol--the RAMESES II study: developing guidance and reporting standards for realist evaluation. BMJ Open. 2015;5(8):e008567.

15. Agency. EM. Guideline for good clinical practice E6(R2). London: Committee for Human Medicinal Products. 2016.

16. J T. Spirit checklist. Cambridge: Harvard Dataverse; 2020.

17. Shams-White MM, Brockton NT, Mitrou P, Romaguera D, Brown S, Bender A, et al. Operationalizing the 2018 World Cancer Research Fund/American Institute for Cancer Research (WCRF/AICR) Cancer Prevention Recommendations: A Standardized Scoring System. Nutrients. 2019;11(7).

18. Moher D, Schulz KF, Altman DG. The CONSORT statement: revised recommendations for improving the quality of reports of parallel-group randomised trials. Lancet. 2001;357(9263):1191–4.

19. Saab MM, Landers M, Cooke E, Murphy D, Hegarty J. Feasibility and usability of a virtual reality intervention to enhance men’s awareness of testicular disorders (E-MAT). Virtual Reality. 2019;23(2):169–78.

20. Gamper EM, Musoro JZ, Coens C, Stelmes JJ, Falato C, Groenvold M, et al. Minimally important differences for the EORTC QLQ-C30 in prostate cancer clinical trials. BMC Cancer. 2021;21(1):1083.

21. Devlin N, Finch AP, Parkin D. Guidance to Users of EQ-5D-5L Value Sets. In: Devlin N, Roudijk B, Ludwig K, editors. Value Sets for EQ-5D-5L: A Compendium, Comparative Review & User Guide. Cham (CH): Springer Copyright 2022, The Author(s). 2022. p. 213–33.

22. Sousa VD, Zauszniewski JA, Bergquist-Beringer S, Musil CM, Neese JB, Jaber AF. Reliability, validity and factor structure of the Appraisal of Self-Care Agency Scale-Revised (ASAS-R). J Eval Clin Pract. 2010;16(6):1031–40.

23. Saab MM, McCarthy M, Davoren MP, Shiely F, Harrington JM, Shorter GW, et al. Enhancing Men’s Awareness of Testicular Diseases (E-MAT) using virtual reality: A randomised pilot feasibility study and mixed method process evaluation. PLoS One. 2024;19(7):e0307426.

24. Viechtbauer W, Smits L, Kotz D, Bude L, Spigt M, Serroyen J, et al. A simple formula for the calculation of sample size in pilot studies. J Clin Epidemiol. 2015;68(11):1375–9.

25. Braun VC V. Thematic Analysis: A Practical Guide: SAGE; 2022.

26. Eldridge SM, Chan CL, Campbell MJ, Bond CM, Hopewell S, Thabane L, et al. CONSORT 2010 statement: extension to randomised pilot and feasibility trials. BMJ. 2016;355:i5239.

27. O’Connor MD, F; O’Donovan, B; Donnelly, C. The Unmet Needs of Cancer Survivors in Ireland: A Scoping Review 2019. https://www.ncri.ie/sites/ncri/files/pubs/HSE%20Report%203%20-%20Unmet%20needs%20of%20cancer%20survivors%20in%20Ireland%20Final%20Version.pdf: National Cancer Registry Ireland; 2019.

